# Sleep quality in the chronic stage of concussion is associated with poorer recovery: A systematic review

**DOI:** 10.1101/2020.09.04.20188425

**Authors:** Rebecca Ludwig, Eryen Nelson, Prasanna Vaduvathiriyan, Michael A. Rippee, Catherine Siengsukon

**Author notes:** **Corresponding Author**: Catherine Siengsukon, PT, PhD University of Kansas Medical Center Physical Therapy and Rehabilitation Sciences 3901 Rainbow Blvd, Mailstop 2002.

## Abstract

**Objective:** To examine the association between sleep quality during the chronic stage of concussion and post-concussion outcomes.

**Literature Survey:** Literature searches were performed during July 1^st^ to August 1^st^, 2019 in selected databases along with searching grey literature. Out of the 733 results, 702 references were reviewed after duplicate removal.

**Methodology:** Three reviewers independently reviewed and consented on abstracts meeting eligibility criteria (n = 35). The full-text articles were assessed independently by two reviewers. Consensus was achieved, leaving four articles. Relevant data from each study was extracted using a standard data-extraction table. Quality appraisal was conducted to assess potential bias and the quality of articles.

**Synthesis:** One study included children (18–60 months) and three studies included adolescents and/or adults (ranging 12 years to 35). The association between sleep and cognition (two studies), physical activity (one study), and emotion symptoms (one study) was examined. Sleep quality was associated with decreased cognition and emotional symptoms, but not with meeting physical activity guidelines 6 months post-concussion injury.

**Conclusions:** The heterogeneity in age of participants and outcomes across studies and limited number of included studies made interpretations difficult. Future studies may consider if addressing sleep quality following concussion will improve outcomes.

## Introduction

The prevalence of a concussion is 1.7- 3.8 million people each year in the United States.^1#x2013;3^ Concussion injuries occur in a variety of settings including motor vehicle accidents, sports, recreation, falls, work environments, and war blast zones^4,5^ and are commonly a result of a high magnitude of force placed to the head, neck, or face.^2,6^ Symptoms caused by a concussion are often categorized into four domains: somatic, cognitive, mood disruption, and sleep dysregulation.^3,7^ *Somatic symptoms* include vertigo, sensitivity to light and sound, nausea, vomiting, headaches, and migraines.^7^ Slower processing speed, decreased focus, disrupted short term memory, and difficulty concentrating are commonly reported impairments in *cognition* following a concussion.^7^ *Mood disruptions* after a concussion consist of an increase or onset of anxiety, depression, or irritability.^7^ *Sleep dysregulation* often manifests in the form of hypersomnia, insomnia, or circadian rhythm disruption.^7^ Acutely following the injury, headache, fatigue, dizziness, and difficulty with information processing are the most endorsed symptoms.^8^,^9^ Symptoms that are commonly reported at follow-up visits are sleep disturbances, frustration, forgetfulness, and fatigue.^8^

Recovery time frames after the concussion injury are different for each individual and commonly are grouped in the following stages: *acute stage* (the first seven days), *sub-acute stage* (8-89 days), and *chronic stage* (≥ 90 days) following the concussion injury.^10^ Symptoms resolve for 80-90% of individuals 1-3 weeks post-injury.^1,4,6,7^ However, for the remaining 10–20% of individuals, symptoms persist for months to years post injury.^11–14^

While symptom severity and duration are difficult to predict,^15^ there are certain factors from prior to the injury, the injury itself, and after the injury that increase the risk of prolonged symptoms. Pre-injury risk factors include age (0-4 years old and 65+ years old), history of learning disabilities, migraines, previous concussions, psychiatric conditions, female gender, lower cognitive reserve, lower socioeconomic status, substance abuse, and premorbid sleep disturbance.^16–20^ Concussion-specific risk factors for prolonged symptoms consist of loss of consciousness, injuries to other areas of the body, lower (< 15) Glasgow Coma score, and a higher velocity mechanism of injury.^16,17^ Post-injury anxiety, depression, post-traumatic stress disorder, migraine, sleep disturbances, fogginess, difficulty concentrating, vestibular dysfunction, and ocular motor dysfunction post-injury have also been associated with prolonged symptom recovery.^13,14,21^ Variability in length of recovery following a concussion is likely due to multiple reasons, including fluctuation of cellular effects, extent of axonal injury, location of impact, and the number and severity of symptoms following a concussion.^22–25^ Research is still investigating why various risk factors and combinations of risk factors contribute to delayed concussion recovery.

Sleep disturbance prior to concussion as well as the development of sleep disturbances following a concussion have also been associated with prolonged concussion symptoms. Poor sleep quality is related to increased pain,^26^ more frequent headaches,^27^ and vestibular issues.^28^ Our recently published systematic review found that sleep disturbances in the acute phase have been shown to be predictive of prolonged symptom recovery following a concussion.^17^ Short sleep duration and poor quality during the acute and subacute stage of concussion are associated with a range of prolonged post-concussion symptoms including poorer cognitive function, reduced productivity at work or school, decreased social engagement, increased pain, daytime sleepiness, depression, and anxiety.^17,29,30^

The association between sleep and post-concussion symptoms in the acute and sub-acute stages has been well documented.^19,31-36^ There remains a gap in the literature assessing the association between sleep and post-concussion symptoms in the chronic phase of concussion recovery. Therefore, the purpose of this systematic review is to examine the association between sleep disturbance and post-concussive symptoms at > 6 months post-injury.

## Methods

To conduct this systematic review, literature searches were performed between July 1^st^ to August 1^st^, 2019 in the following databases: Ovid MEDLINE(R), Cumulative Index of Nursing & Allied Health Literature (CINAHL), and Web of Science - Science Citation Index Expanded. relevant cited references and grey literature including clinical trials and conference papers were searched. The review was limited to studies published in the English language using the databases’ language limit feature. Publication date limits to the search were not applied.

A combination of keywords and controlled vocabulary terms were used in Medline and CINAHL to execute the searches in the databases. Medical subject headings including “brain concussion”, “brain injuries”, “traumatic brain injuries”, “sleep”, “pain”, “pain management”, and keywords (text-words) such as “mild traumatic injuries”, “concussion”, “post-concussion” were used in various combinations to increase recall.

PRISMA guidelines were followed for the article selection. Studies included in the systematic review, had to meet the following inclusion criteria: (1) included people with concussive or mild traumatic brain injury; (2) assessed sleep using subjective or objective measures > 6 months post-concussion; (3) used a prospective or cross-sectional study design; (4) reported other non-sleep outcomes > 6 months post-concussion; and (5) included statistical analyses to relate sleep outcomes to other reported outcome(s). Studies were excluded if they: (1) included animals; (2) included participants with moderate and/or severe traumatic brain injury;

(3) included participants with other neurologic conditions; (4) used a study design other than prospective or cross-sectional; (5) were irrelevant to the objective of the systematic review; (6) were published in a language other than English; or (7) included a duplicate data set.

Three reviewers (R.L., K.S., A.P.) assessed titles and abstracts of all potentially eligible articles for inclusion/exclusion criteria to identify which titles/abstracts met those criteria and would be considered for the systematic review. The full-text article of all considered studies was then retrieved and reviewed independently by two reviewers (R.L., E.N.) to establish article relevance. The reviewers came to a consensus on which abstracts met the inclusion/exclusion criteria and which full-text articles were included in the systematic review.

### Data Extraction

Relevant data were extracted from each included study by two reviewers (R.L., E.N.) using a data-extraction table. Extracted information included the author(s), year published, study location, study design, sample size, participant demographic characteristics, time since injury, measures used to assess sleep, measures used to assess other outcomes, and results.

### Quality Assessment

The risk of potential bias as well as the overall quality of the included studies was assessed using the *Joanna Briggs Institute Critical Appraisal Checklist for Analytical Cross Sectional Studies*.^37^ The checklist is comprised of eight questions regarding defined study inclusion criteria, description of study participants, exposure validity and reliability, standard criteria for measurement conditions, identification of confounding factors, strategies for dealing with confounding factors, valid and reliable outcomes measured, and appropriate statistical analysis. All articles were scored independently by two reviewers (R.L., E.N.) as “yes”, “no”, “unclear”, or “not applicable” for each item on the checklist. The reviewers met to reach consensus.

## Results

The search strategy generated a total of 733 articles. There were 31 duplicates removed, leaving 702 titles/abstracts to review. After the review of title/abstracts for inclusion/exclusion criteria, 662 articles were removed. A total of 40 full-text articles were reviewed. Thirty-six articles were removed due to: reasons related to unclear assessment time since injury (n = 22), no statistical analyses conducted to determine the association between sleep and other post-concussion outcomes (n = 11), and inability to locate the full-text article (n = 3). A total of four articles met the criteria to be included in this review (Figure 1).

**Figure 1.**
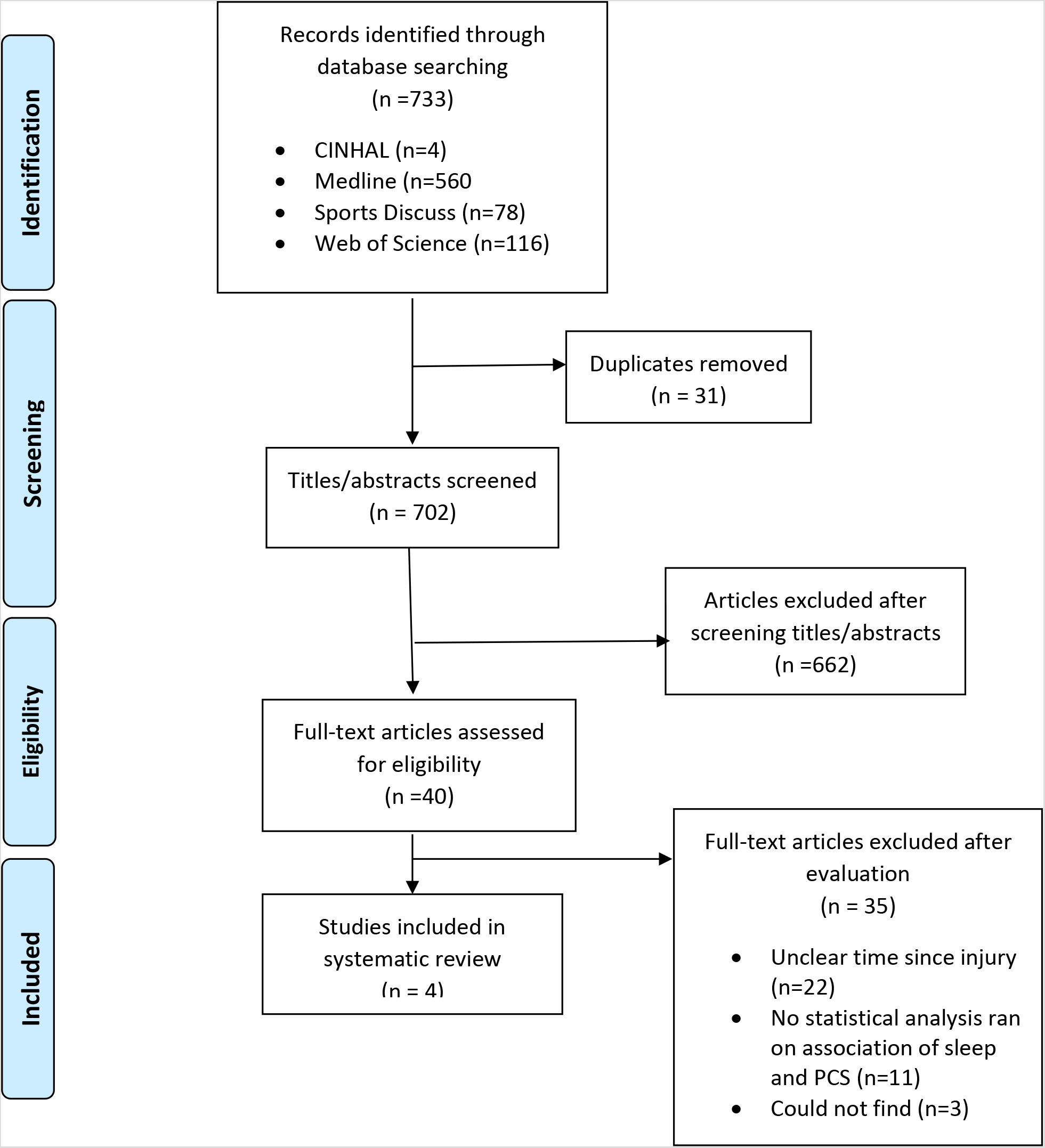

The studies in this review are summarized in the data extraction table (Table 1) and the risk of bias is reported in the quality appraisal table (Table 2). The number of participants in each study ranged from 49-232 individuals. One article included pediatric patients (18-60 months)^38^ and three articles included adolescents and adults (>12 years old).^39–41^

**Table 1.**
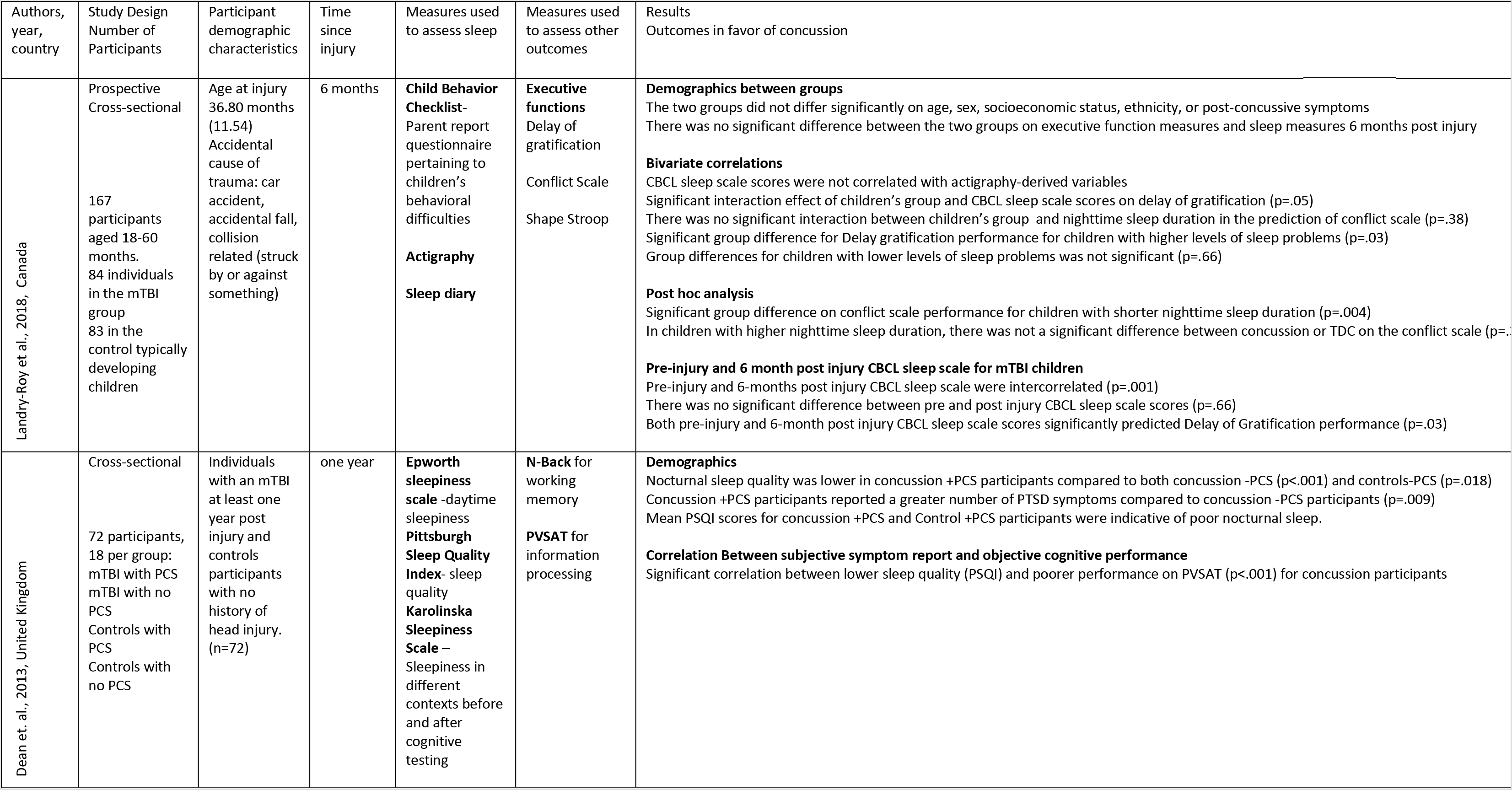
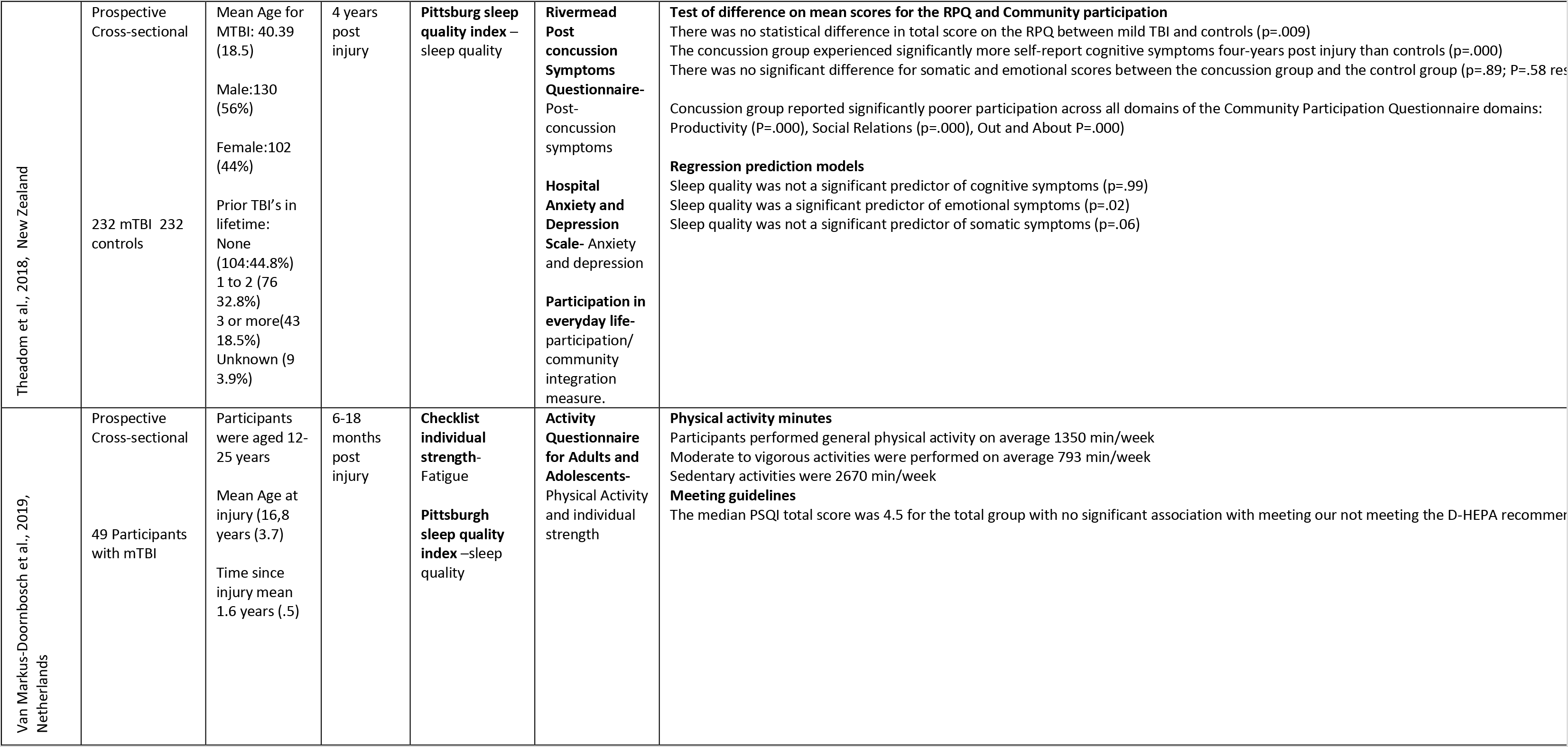
Results

**Table 2.** Bias Table

| Authors (year) | Criteria for inclusion clearly defined | Participants described in detail | Exposure measured in valid and reliable way | Objective standard criteria used for measurement | Confounding factors identified | Strategies to deal with confounding factors | Outcomes measured in a valid and reliable way | Appropriate statistical analysis used |
| --- | --- | --- | --- | --- | --- | --- | --- | --- |
| Landry-roy et al 2018 | Yes- defined inclusion/ exclusion criteria | Yes- gave specific ages, and criteria for meeting concussion standards | Yes- concussion was evaluated and defined in a valid reliable way | Yes- used standard, valid, and reliable assessment tools | No- did not identify confounding factors | No- did not control for confounding factors in their statistical analysis | Unsure- Use of the CBCL sleep portion may not be specific enough | Unsure- did not describe in detail how the statistical analysis answered their research question |
| Dean and Sterr, 2013 | Yes- defined inclusion/ exclusion criteria | Yes- Gave criteria for meeting concussion standards | Yes- concussion was evaluated and defined in a valid reliable way | Yes- used standard, valid, and reliable assessment tools | No- did not identify confounding factors | Unsure- Did not identify confounding factors and unclear if they statistically controlled for factors. | No- the researchers allowed for rest breaks and potential days between assessment times | Yes- used appropriate statistical analysis |
| Thea.dom et al., 2018 | Yes- defined inclusion/ exclusion criteria | Yes- gave criteria for meeting concussion standard | Yes- concussion was evaluated and defined in a valid reliable way | Yes- used standard, valid, and reliable assessment tools | Yes- identified confound factors | Yes- controlled for confounding factors in statistical analysis | Yes- met in homes or over phone, done by trained research personnel | Yes- used appropriate statistical analysis |
| Markus- Doornbosch et al., 2018 | Yes- defined inclusion/ exclusion criteria | Yes – gave criteria for meeting concussion standard | Yes- concussion was evaluated and defined in a valid reliable way | Yes- used standard, valid, and reliable assessment tools | Yes- identified confounding factors | Not Applicable | Unsure | Yes- used appropriate statistical analysis |

Landry-Roy et al. (2018)^38^ evaluated the relationship between sleep and executive function in preschool children. Children aged 18-60 months (n = 167) who either had a concussion (n = 84) or were typically developing children (TDC) (n = 83) participated. The results indicated there was a significant interaction of children’s group (concussion or TDC) and the Children Behavior Checklist (CBCL) sleep scores on the Delay of Gratification assessment (p = .05) at 6-months. Suggesting that children with a concussion and sleep disturbance have disrupted executive function. There was a significant group difference for the Delay of Gratification assessment performance for children with higher levels of sleep problems on the CBCL (p = .03). Children with a concussion had decreased performance compared to TDC on the delay of gratification if they had higher levels of sleep problems on the CBCL. Similarly, in a post hoc analysis there was a significant group difference on Conflict Scale performance for children with shorter sleep duration (p = .004). Thus, children with a concussion exhibited poorer conflict scale performance than TDC only if they had shorter nighttime sleep duration. There was a small subgroup of children (n = 68; 33 concussion 35 TDC) who wore actigraphy but no association between actigraphy and sleep diary and executive function was found. The results of this study indicate that the combination of a concussion and poor sleep result in poorer executive functions in pre-school aged children.

The study by Dean and Sterr (2013)^39^ evaluated working memory and information processing speed in adults (n = 36; 20-30 years of age) at least one-year post concussion injury compared to a non-head injury control group (n = 36; 20-30 years of age). Participants were divided into one of four groups: 1. Concussion + Post-Concussion Syndrome (PCS); 2. Concussion – PCS; 3. Control (non-head injury) + PCS; 4. Control (non-head injury) - PCS. Nighttime sleep quality was lower in the Concussion + PCS participants compared to the Concussion - PCS and Control – PCS (p = .018). Mean Pittsburg Sleep Quality Index (PSQI) scores for the Concussion + PCS (8.6 +/− .8) and the Control + PCS (6.6 +/−.7) were indicative of poor nighttime sleep. There was a significant correlation for all individuals with a concussion between lower PSQI scores and poorer performance on the N Back (working memory) and Paced Visual Serial Addition Task (information processing speed) (Rho = .62, p<.001).

Theadom et al. (2018)^40^ evaluated how sleep predicts post-concussion symptoms and community and social engagements in adolescents and adults (n = 232; age ≥ 16) from the population based BIONIC study^31,32^ four years after the initial concussion injury. The study used the Rivermead Post Concussion Symptoms Questionnaire to assess post-concussion symptoms and the PSQI for sleep quality. The concussion group reported significantly more cognitive symptoms four years post-injury compared to controls. There was no difference between the concussion and control groups on somatic and emotional scores. Regression modeling revealed that sleep quality was a significant predictor for emotional symptoms on the Rivermead Post Concussion symptom questionnaire (p = .02) but not for cognitive (p = .99) or somatic symptoms (p = .06). In particular, poorer sleep quality was a predictor of higher anxiety, depression, and irritable mood symptoms 4 years following concussion.

Van Markus-Doornbosch et al. (2019)^41^ evaluated the relationship between sleep quality and physical activity level in adolescents and adults who had a concussion. A total of 49 participants (12-25 years old) who had a concussion 6-18 months prior were enrolled in the study. If the participant was < 18 years old, a parent completed the questionnaires. Physical activity was assessed by the Activity Questionnaire for Adults and Adolescents, and sleep quality was assessed using the PSQI. Twenty-five out of 49 (51%) participants did not meet the Dutch Health Enhancing Physical Activity (D-HEPA) recommendations. The median PSQI total score was 4.5 for the entire study group with no significant association with meeting or not meeting the D-HEPA recommendations (OR .99, 95% CI .82,1.19).

The two contributing factors for bias were not adequately controlling for confounding factors and not using an appropriate assessment tool for measuring sleep outcomes (Table 2). In two out of the four studies, the confounding factors were not adequately described or controlled for in the statistical analysis. One out of the four studies did not use an assessment that was specific to assess sleep outcomes. The sleep portion of the CBCL used in Landry-Roy et al. has good convergent validity, but there is debate on if this assessment is specific to sleep when compared to other pediatric sleep assessments.

## Discussion

This systematic review found that poor sleep quality is associated with concussion symptoms 6 months to 4 years following the injury. Poor sleep quality was associated with impaired executive function in preschoolers at 6 months following concussion,^38^ and working memory and information processing speed in adults.^39^ Poor sleep quality was also found to be a predictor of emotional symptoms in adults 4 years post-concussive injury.^40^ However, one study did not find a significant association between sleep quality and meeting physical activity guidelines in adolescents.^41^ Only four articles met the criteria for inclusion in this systematic review, thus limiting the scope of interpretation, while simultaneously highlighting the lack of research on how poor sleep quality and sleep disturbances are associated with prolonged symptoms in individuals with chronic concussion.

Two articles in this systematic review examined the association between sleep and executive function, working memory, and information processing speed.^38,39^ Cognitive function has been shown to be impaired in the acute and subacute stages of concussion.^14,16,42^ Symptoms such as taking longer to think, decreased focus, and decreased processing speed are commonly reported in the acute and subacute stages.^14,16,42^ What was observed from this systematic review was that executive function, working memory, and processing speed was still impaired.^38,39^ Sleep disturbance and cognitive disturbance are ranked as the most common complaints of patients at follow-up visits.^16^ The association between sleep disturbance and cognitive disturbance is not a surprise as sleep disturbance can influence cognition.^43^ Individuals recovering from a TBI have shown sleep fragmentation and altered sleep architecture on polysomnograpy.^44–46^ More specifically, individuals recovering from a TBI spend more time in slow wave sleep (SWS) and less time in rapid eye movement (REM) sleep.^44^ While SWS is necessary for cellular and structural recovery, REM is needed for information processing and memory consolidation. Therefore, a reduction in REM could contribute to impairments of executive functioning, working memory, and information processing that are often experienced by individuals following concussion.^47–51^ Psychological factors such as depression, anxiety, and mood influence cognition as well.^52–54^ Sleep disturbances and psychological factors are interrelated, so it is likely a combination of sleep disturbance and psychological factors impact cognitive function following a concussion^55^

Sleep quality was also found to be predictive of emotional symptoms including anxiety, depression, and irritability 4 years following a concussion.^40^ This finding supports evidence that chronic poor sleep and insomnia can contribute to the development of depression, anxiety,irritability.^56–60^ Furthermore, screening and treating insomnia reduces concomitant symptoms of depression, anxiety, and stress.^61–63^ While sleep quality and insomnia contributes to emotional symptoms, research also shows that emotional symptoms can contribute to the development of sleep issues.^64^ A negative mood (anxious, stressed, tense, sad, or irritable) is more common in individuals with poor sleep.^65,66^ There is a strong relationship between the development of anxiety and the development of poor sleep and insomnia as many core features (intense worry or fear, and avoidance behavior) are the same.^67^ Health care providers should screen for both sleep and emotional issues in people following concussion, and more studies are needed to determine optimal prioritization of treatment for sleep and emotional issues.

One study found that meeting physical activity guidelines was not associated with sleep quality six months post-injury.^41^ This is surprising as physical activity and sleep have been shown to have a reciprocal relationship.^68^ If an individual does not have a quality night of sleep, there is decreased motivation to be physically active the following day.^68^ Also, physical activity improves sleep duration and quality by increasing sleep drive.^69^ The lack of association in Van Markus-Doornbosch^41^ might be due to the mean PSQI score was 4.5, indicating the participations on average had good quality sleep. Also, the authors assessed the relationship between sleep quality and adherence to physical activity guidelines, not physical activity specifically. Van Markus-Doornbosch^41^ reported that within their sample 51% of individuals following concussion met physical activity guidelines which is comparable to the national average in the United States. For adults in the United States, 44.8% of individuals meet physical activity guidelines.^70^ Therefore, it is questionable on if guidelines for physical activity matter when assessing sleep in individuals with a concussion.

The limitations within the articles for this systematic review is due to not controlling for confounding factors and the specific sleep assessments used. Within the studies reviewed two out of the four articles did not describe confounding factors or address them within their statistical analysis. This made interpretation of the results challenging as it is unclear what effect the confounding factors had on the results described. Additionally, the sleep portion of the CBCL used in Landry-Roy et al., 2018 had good convergent validity, but it is of question on if this assessment is specific enough to assess sleep in comparison to other pediatric sleep questionnaires.

This systematic review does not come without limitations. One limitation is there are only four articles included within this review, limiting the breadth and depth for interpretation. Additionally, the studies included in this systematic review includes a variety of outcomes. The heterogeneity limits interpretation as each study addresses different outcomes (cognition, physical activity, and emotional symptoms). There needs to be more studies evaluating these outcomes to strengthen the knowledge base on how sleep affects cognition, physical activity, and emotional symptoms.

This systematic review indicates that poor sleep quality is associated with impairment in executive function,^38^ working memory, information processing,^39^ and emotional symptoms,^40^ but not physical activity in individuals > 6 months post-concussion. Additional studies are needed to more thoroughly understand how sleep is associated with various post-concussion symptoms in the chronic stage. Also, investigating how addressing sleep issues may impact symptom resolution and recovery in people with a concussion is warranted.

## Data Availability

No data was taken for this systematic review

## Acknowledgements

Kelli Stallbaumer and Alexandra Ptacek assisted with screening abstracts for full text review. This paper is partially funded through the T-32 neurologic rehabilitation training grant through the University of Kansas Medical Center.

## Notes

### Competing Interest Statement

The authors have declared no competing interest.

### Funding Statement

This review is funded through the T-32 training grant at the university of Kansas medical center.

### Author Declarations

No IRB needed for systematic review

